# Part-time or full-time teleworking? A systematic review of the psychosocial risk factors of telework from home

**DOI:** 10.1101/2022.07.22.22277922

**Authors:** Evelise Dias Antunes, Leonardo Rodrigues Thomaz Bridi, Marta Santos, Frida Marina Fischer

## Abstract

Since the start of the Coronavirus pandemic thousands of people have experienced teleworking and this practice is becoming increasingly commonplace. Systematic reviews can yield evidence and information to help inform the development of policies and regulations, the aim of this study was to highlight the differences in exposure to psychosocial risk factors for health between part-time and full-time teleworking from home. The protocol of the systematic review of the literature was registered on PROSPERO 2020 platform - International Prospective Register of Systematic Reviews (number CRD42020191455), according to the PRISMA statement guidelines. The key words “telework” and frequency (“part-time” or “full-time”), together with their synonyms and variations, were searched. Independent researchers conducted the systematic search of 7 databases: Scopus, SciELO, PePSIC; PsycInfo, PubMed, Applied Social Sciences Index and Abstracts (ASSIA) and Web of Science. Of the 638 articles identified from 2010 to June 2021, 32 were selected for data extraction. The authors evaluated the risk of bias and quality of evidence of the studies included using the Mixed Methods Appraisal Tool. The results were categorized into 7 dimensions of psychosocial risk factors: work intensity and working hours; emotional demands; autonomy; social relationships at work; conflict of values, work insecurity and home/work interface. The results revealed scant practice of full-time teleworking prior to the pandemic. Regarding the psychosocial risk factors found, differences were evident before and during the COVID-19 pandemic. For part-time and full-time telework prior to the pandemic, the dimensions of intensification of work and working hours, social relationships at work, and the home-work interface were the most prominent factors. However, in studies performed during the COVID-19 pandemic where teleworking was mostly performed full-time, there was an increase in focus on emotional demands and the home-work interface, and a reduction in the other dimensions.

## 1 INTRODUCTION

Teleworking is a reality for millions of employees worldwide. The nature of telework can vary from compulsory to being a consequence of new ways of working. During the pandemic, this approach constituted a solution to minimize economic impact and maintain some activities and functions conducive to continuation through the use of information and communication technologies (ICT). It was originally thought that teleworking would be temporary, i.e. required only until a sufficient contingent of the population had been vaccinated to allow a return to the workplace. However, in many cases, the result was a structural change in the way of working.

Experiences differed depending on country, region, type and organization of work, residence, family composition, among other factors. For companies, there were clear benefits in terms of savings in overheads such as rent, water, electricity, telephone, internet, cleaning, maintenance, consumables, besides reduced transport costs with commuting of workers.

For employees, benefits appear to be offset with drawbacks, with some individuals in favor but also some inquiring. There is mixed opinion, with some not wishing to telework voluntarily, while others intend to continue the practice part-time or full-time. Consequently, there has been much debate over the return to face-to-face activities and continuation of teleworking. Based on the premise that differences exist between part-time and full-time teleworking, we posed the research question “what are the differences in psychosocial risk factors between part and full-time teleworking?”.

Teleworking is not a new area in the literature. The precursor of studies on this modality was conducted by the North American Jack Nilles, investigating the potential impact of the use of telecommunications and computer technology as an alternative to urban transportation, creating workstations at centers close to home or within them, thereby dispensing with the need to commute to work [1–5].

In 1985, Niles [6] predicted that, within a decade, 5 million white-collar workers would be able to work from home. He identified a number of possible barriers to acceptance of telecommuting by workers: fear of change by workers, the new technology, personal isolation, interruption in career, threat to job security and worker exploitation. Reasons among employers were more varied: concerns of decline in productivity, increase in turnover rates, a reduced ability to adapt to changes, the legal issues and their regulation, safeguards against exploitation and the physical safety of employees working from home [6].

Over previous decades, despite the technology revolution and globalization, teleworking never took off in the way it has today. This modality existed before the pandemic, but was available to a relatively small contingent of workers. Participation in teleworking has tripled since the start of the pandemic [7]. For example, in the European Union, the proportion of teleworkers rose from 11% to 48% during the pandemic [8, 9]. In Latin America and the Caribbean, over 23 million people had transitioned to teleworking in the second quarter of 2020 [10].

Following the pandemic, the full-time modality was adopted, even in job areas hitherto regarded as unfeasible, such as telemedicine. Two years on, the question is whether teleworking is set to become the “new normal”. Thus, before assuming this will be the model of working for the future, it is important to elucidate the impacts on health of those engaged in teleworking.

Unlike some technology firms such as Twitter, which allows its staff to telework from anywhere in the world, some large companies have decided the opposite, calling on workers to return to the office in 2022 [11].

However, not so long ago, major companies such as Yahoo and Bank of America had already discontinued full-time teleworking. The technology firm Yahoo ordered home workers back to the office, detailing in a memo the shift in policy of the companýs Human Resources Department, stating that face-to-face interaction between workers promotes a more collaborative culture [12].

Therefore, the present review was prompted by the fact that thousands of people have experienced teleworking since the start of the Coronavirus pandemic and given the tendency of this practice to become more commonplace in the near future. There is also an apparent acceleration of the political agenda in many countries toward regulating this work modality [13]. Given that systematic reviews can yield evidence and information to help inform the development of policies and regulations, this study aimed to highlight the differences between the psychosocial risk factors comparing part and full-time teleworking from home.

Among the previous available systematic reviews on teleworking, no differentiation was made between the place and frequency basis of teleworking, i.e. part and full-time [13–15]. In order to provide supporting background on the topic of teleworking, the ensuing sections outline: 1) Terms, definitions and forms, organizations of telework; and 2) Psychosocial factors in teleworking and the model used as a conceptual framework for organizing and guiding the discussion of the literature.

### 1.1 Flexible work arrangements to teleworking

Flexible work arrangement (FWA) is a multi-dimensional concept used as an umbrella term to describe new ways of organizing work. Telework, remote work, work at home, home-based work, telecommuting, remote work from home, e-work and smart work are terms found in the literature referring to FWAs [16]. Although related and with some degree of overlap, these terms represent distinct situations. They can refer to flexible hours, such as flextime, and/or flexibility in the workplace, such as teleworking, flexiplace and e-working [14].

Defining these concepts to measure them has become more important than ever since the massive lockdown imposed by the pandemic. The lack of statistical standards defining conceptual differences poses a challenge, because this impacts the data, and consequently, international comparative studies [17].

There is currently no internationally standardized conceptual definition and ways of measuring the incidence of remote working or teleworking, although they are associated with flexibility in terms of working time and place of work [17, 18].

Remote work can be described as situations in which the work is performed, partially or fully, at a place outside of the standard work premises (off-site). These alternative workplaces range from co-working spaces to cafes, bookshops, and client premises. In addition, workers can be independent (self-employed) or employees. However, workers of family businesses that do not have a fixed workplace, and also independent workers, are typically excluded from this remote work group [17].

Telework is a unique category in which workers must use information and communication technologies (ICT) to perform remote working [16]. It should be noted that the joint study on teleworking carried out by the ILO and Eurofound was restricted to employees only [18].

Home-based workers constitute those for whom their main place of work is their home, representing a subcategory of work at home. Working predominantly from home differs to working out of different places (public spaces, co-working, client premises), and akin to frequency, the use of work spaces are aspects which, according to intensity, may have different consequences for working conditions [18].

In the present review, the term part-time telework from home is used to refer to home teleworking performed by workers that work from home for just 1 or 2 days a week for example. By contrast, the term full-time telework from home refers to weekly teleworking from home, with occasional visits to the office. Given that teleworking may be full or part-time, it is important to explore differences between work arrangements, the impact on the work-life balance, working hours, productivity, health and well-being [19]. It follows that the intensity of teleworking should also be taken into account in field research, as opposed to merely comparing teleworkers versus non-teleworkers [20–23].

### 1.2 Conceptual framework

This section presents the conceptual framework underpinning the discussion, outlining the concepts and dimensions of the psychosocial risk factors covered by the study. The COVID-19 pandemic was accompanied by an increase in the inequalities that already existed in teleworking and an exacerbation of the risks associated with the practice [24]. Given the importance of teleworking during 2020 and 2021, and its continuity in many companies despite the significant decline in the number of COVID-19 cases, attention has turned to the psychosocial factors associated with teleworking.

As is the case of teleworking, psychosocial risk factors for health in the workplace are not a new phenomenon. In 1984, the International Labor Organization (ILO) [25] published the first official document with this issue on its agenda, alerting to the growing incidence and need for intervention and programs to protect workerś health. In the document, psychosocial factors at work were defined as “interactions between and among work environment, job content, organizational conditions and workers’ capacities, needs, culture, personal extra-job considerations that may, through perceptions and experience, influence health, work performance and job satisfaction” [25] (p.3).

Since then, decades of studies have been carried out on this area, yet with no consensus on interpretation under different conceptual models [26, 27]. Therefore, the conceptual framework of Gollac and Bodier [28] was adopted for this study, given that it was derived from a report produced by a panel of experts set up upon request of the French minister for work, tasked with monitoring psychosocial risks at work.

According to Gollac and Bodier [28], psychosocial factors are “risks to mental, physical and social health caused by working conditions and by organizational and relational factors subject to interaction with mental functioning” (p.13). These authors grouped the factors into 6 dimensions:

1. Intensity of work and working hours
2. Emotional demands
3. Autonomy
4. Social relationships at work
5. Conflicts of values
6. Job insecurity

This classification seeks to understand how the different dimensions of work influence and/or determine the process of illness. This framework was used to group and discuss markers of psychosocial risk from the European Working Conditions Survey (EWCS), employed to monitor the countries of the European Union [29].

For the first dimension addressing intensity of work and working hours, the immediate consequences/determinants of intensity and complexity of work must be examined, along with the duration and organization of work time [28].

Emotional demands, on the other hand, are connected with relations with the public and can be particularly high for those who work with suffering or pain; these can be positive, but may also cause distress [30].

Autonomy denotes the possibility of the worker having control over their work through involvement with the output. The risk related to this aspect lies in a lack of autonomy [28].

The fourth dimension – social relations at work - consists of the social relationships between workers, and also the relationship between worker and the employer. This aspect includes a number of variables, such as opportunities for cooperation; satisfactory support in difficult situations; situation of strain, excessive competition; collective autonomy; participation; technical support received from superiors; human relationships; leadership and style of facilitation; valuing of work; remuneration and career; suitability of the task for the individual; work appraisal; procedural fairness; care regarding wellbeing of workers; recognition by clients and the public; social value of the job; effects of violence; forms of internal violence; and violence at work [28].

Conflict of values refers to ethical suffering when an individual is asked to act in breach of their professional, social or personal values [28].

The sixth and last dimension – job insecurity – includes the socioeconomic insecurity risk of uncontrolled change in the work task and conditions. Socioeconomic insecurity encompasses, for workers, the chance of losing onés job, insecurity over maintaining salary levels or lack of the usual trajectory of career advancement for the job [28].

## 2. METHOD

Based on methodological standards, a systematic review protocol was devised and registered on the PROSPERO platform - International Prospective Register of Systematic Reviews (PROSPERO 2020 CRD42020191455) in June 2020. The protocol followed the Preferred Reporting Items for Systematic Reviews and Meta-Analyses for Protocols 2015 (PRISMA-P 2015) [31].

The process of constructing the guiding question defining descriptors and search mechanisms began based on the PICO strategy. Regarding the meaning of the PICO acronym: (P) refers to the target Population, (I) denotes the aim of the Intervention or area of interest, (C) cover types of intervention or groups for Comparison, and (O) addresses obtaining Outcomes and involves the effects to be achieved by the intervention (author) [31], in this case: “*What are the differences in the psychosocial effects of part-time and full-time teleworking?”*.

### 2.1 Search strategy

The present search strategy was applied similarly to all databases and consisted of one block of keywords intended to cover the various denominations used for “telework” and frequency (“part-time” or “full-time”) as well as its synonyms and variations:

> “telework*” OR “telecommute*” OR “remote work*” OR “work* from home” OR “distributed work*” OR “flexible work*” OR “homework*” OR “virtual work*” OR “virtual office” OR “mobile *work*” OR “e-work” OR “teletrabalho*” OR “trabalho à distância” OR “trabalho de casa” OR “trabalho distribuído” OR “trabalho flexível” OR “trabalho no domicílio” OR “trabalho virtual” OR “escritório virtual” OR “trabalho móvel” OR “teletrabajo*” OR “trabajo a distancia*” OR “trabajo desde casa” OR “trabajo distribuido” OR “trabajo flexible*” OR “trabajo en casa” O “trabajo virtual*” O “oficina virtual” OR “trabajo móvil*”

Based on the search protocol established, two independent researchers conducted a systematic search of 7 electronic databases: Scopus, SciELO, PePSIC; PsycINFO, PubMed, Applied Social Sciences Index and Abstracts (ASSIA) and Web of Science. All searches were limited to articles in relevant psychological, social, management, health and technological scientific journals published from 2010 to June 2021. Peer-reviewed, full publications, written in English, Spanish or Portuguese were eligible. Searches were re-run prior to the final analysis. Unpublished studies were not retrieved. Additionally, the search strategies of relevant systematic reviews were reviewed.

### 2.2 Study participants/population and types

Participants were part-time or full-time teleworking employees, aged 18 years or older. Teleworking was defined as the use of information and communications technologies (ICTs), such as smartphones, tablets, laptops and desktop computers, for the purposes of working from home. The term part-time teleworking was used to refer to regular telework performed by employees working from home, for example, only one or two days a week.

Original, quantitative, qualitative, and mixed methods studies were included in the search according to the inclusion and exclusion criteria shown in Table 1.

**Table 1.** Inclusion and exclusion criteria of systematic review.

| Inclusion criteria | Exclusion criteria |
| --- | --- |
| Participants work remotely from home i.e. spending at least one day of their working time away from their office using ICTs. | Home-based work such as farming or piecework which does not encompass ICT use to enable performance during work activities.<br>Employees who were making use of telework in other places other than their homes, for example, in hotels, coffee bars etc.<br>Reviews or Discussion papers. |
| A broad range of study types were included: cross-sectional studies, longitudinal studies, qualitative research, case reports, quasi-experimental research and meta-analyses. | Narrative literature reviews, non-research letters, editorials, and animal studies. |
| Formal workers | Studies of unemployed participants, self-employed individuals and freelancers.<br>Studies of disabled employees were excluded to ensure that none of the health issues identified were related to employees' disabilities. |

Source: produced by the authors

### 2.3 Data extraction (selection and coding) and synthesis

Two researchers, blinded to each other’s decisions, independently extracted data from all studies. Titles and abstracts, retrieved using the search strategy, were screened for eligibility to identify studies that met the inclusion criteria. Discrepancies were resolved and three researchers came to an agreement on data to be included in the extraction process. Data extraction included Ref ID; First author; year; citation; study aims; sample population; country where study was undertaken; type of study; characteristics of telework; psychosocial effects measured, and main findings. Mendeley software was used to manage the references. Data were stored in an Excel spreadsheet.

The authors evaluated the risk of bias and quality of evidence of the studies included using the Mixed Methods Appraisal Tool (MMAT) Version 2018 [32]. This is a 5-item checklist that yields a score ranging from 0 (no criteria met) to 5 (all criteria met) and can be used across a range of different study designs, such as randomized controlled trials and observational studies. Two authors independently assessed the study quality of the studies included and discrepancies were resolved afterwards. The heterogeneity of the studies included in this review precluded a statistical summary or meta-analysis. A narrative synthesis [33] was produced from the studies included, centering on the differences between the psychosocial effects of part and full-time teleworking.

During the process of reading and extraction, and rechecking of identified and unidentified categories was carried out. The contents present in the articles were assessed and compared across categories related to psychosocial risk factors as per Gollac and Bodier [28] (Table 2). This activity allowed the references to be built which supported the discussion.

**Table 2.**
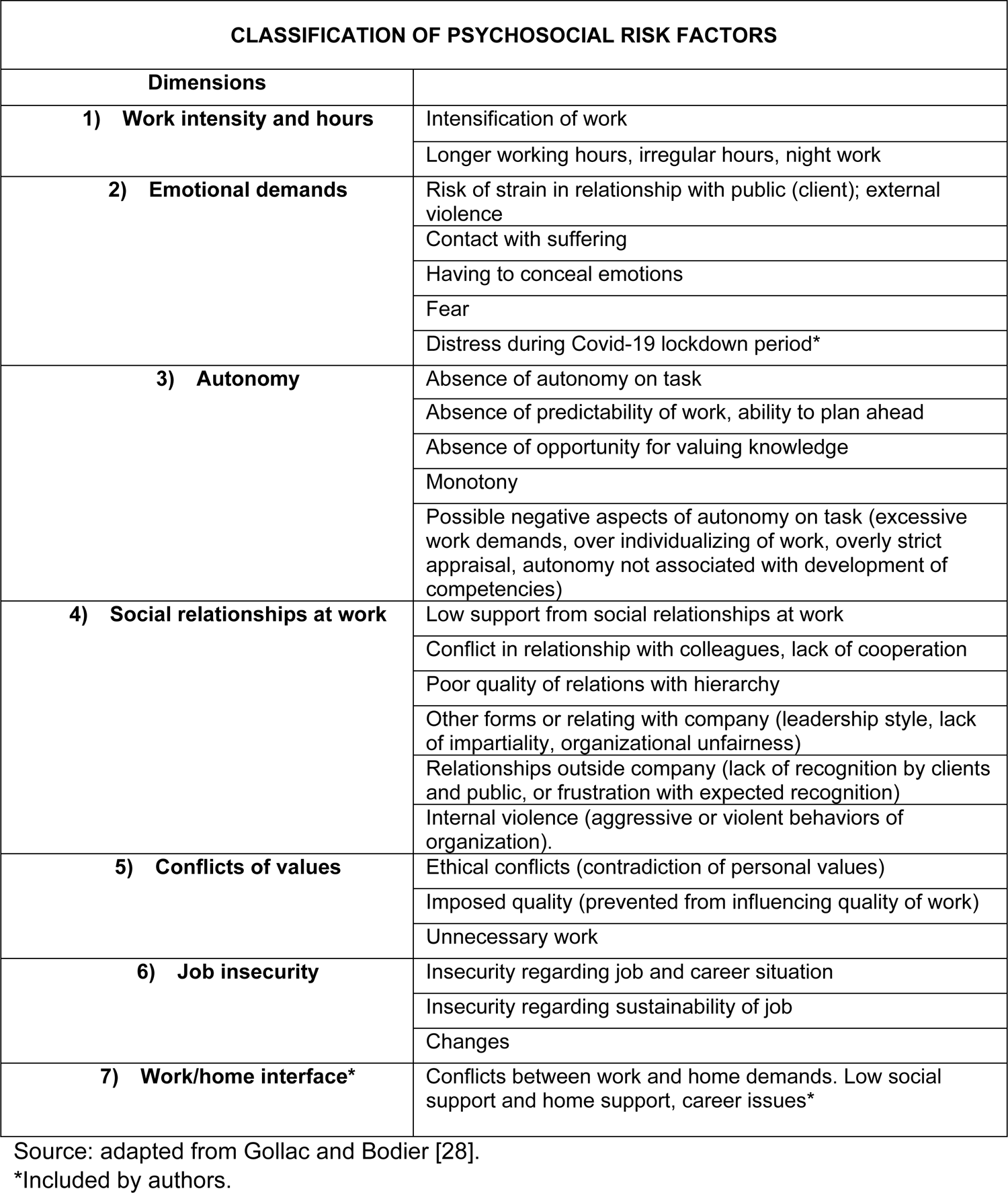
Classification of Psychosocial Risk Factors.

In view of the shift in context during the COVID-19 pandemic and challenges regarding analysis of work, with regard to teleworking, the authors broadened the classification of Gollac and Bordier [28] by including the additional category, 7) *Home/work interface* (to gauge difficulties involving this interface), plus a subcategory associated with emotional demands (category 2), distress during the COVID-19 lockdown period.

Lastly, after classification and analysis of the dimensions, a co-occurrence analysis was performed. Connections and interrelations between the articles and categories under discussion were explored. This comparison was achieved with a map of coded authors and classifications using the MAXQDA Plus 2022 software tool [34].

## 3. RESULTS

According to the PRISMA statement [31], an initial selection was made based on titles and abstracts in order to exclude unavailable sources or publications which failed to meet the screening criteria adopted during the search strategy. Duplicate records due to the multiple databases searched were also removed. Full texts of the selected abstracts were revisited for a more in-depth review. The flow diagram of the PRISMA process summarizing the study selection process is depicted in Fig 1. Data collection took place in two stages, in September 2020 and a later update of more recent articles in June 2021.

**Fig 1.**
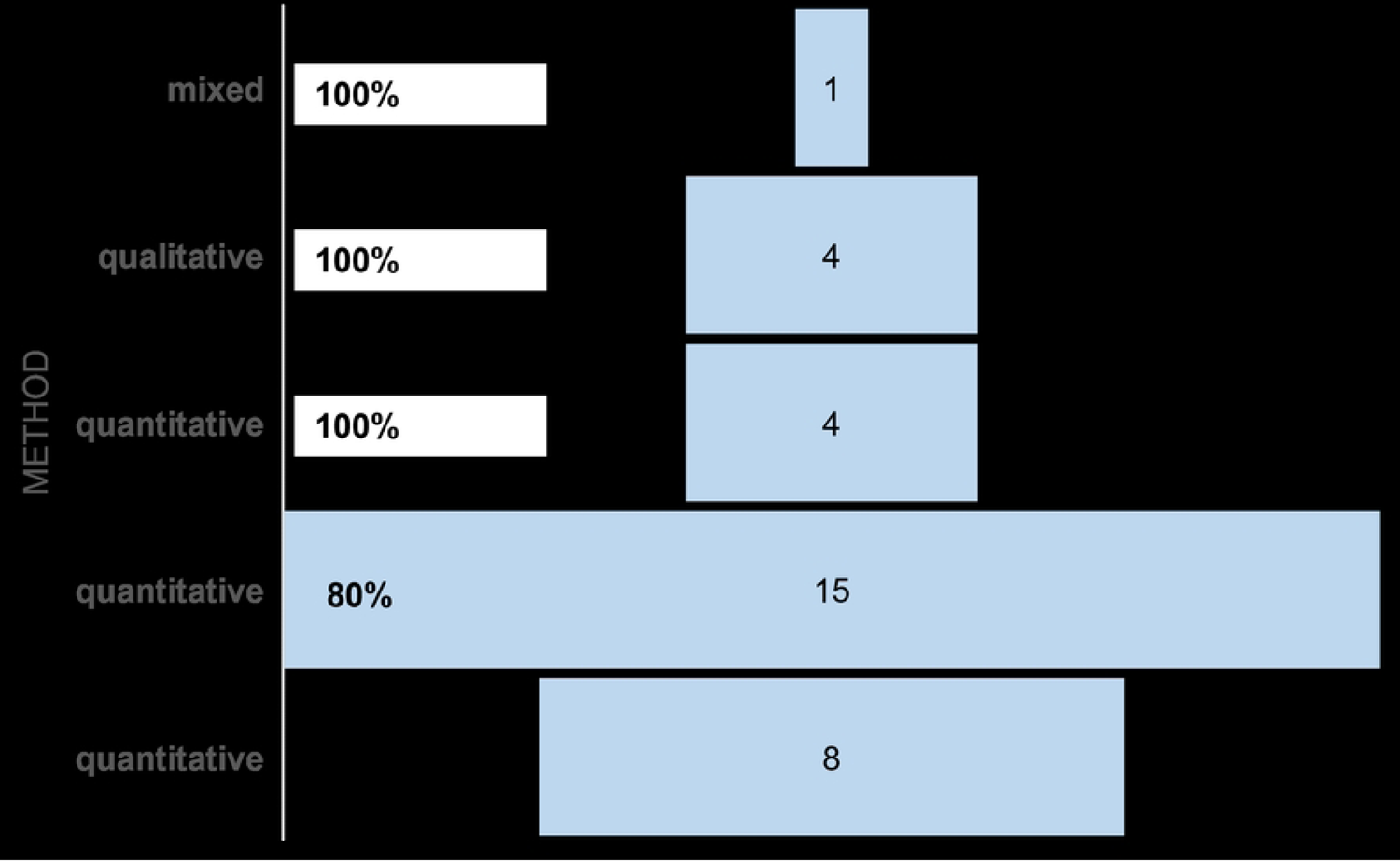
PRISMA Flow Diagram. Source: adapted from Moher *et al* [35]. Source: adapted from Moher *et al* [35].

Of the initial 638 studies retrieved, 114 studies were eligible. A total of 82 articles were excluded at the full-text screening stage because they did not investigate teleworking from home, focusing instead on flexible working practices or flextime work arrangements, or the article content was not related to psychosocial factors, or study had a different type of design to that required (e.g. comments, reviews, published in non-science journals, or were editorials).

During the course of the analysis of the results based on the key words presented in the methods section, the highest number of publications on teleworking was conducted in the United States. However, the wide range of articles found provides a representative view of the subject at least in the Americas, parts of Europe and in some countries of Asia (Fig 2).

**Fig 2.**
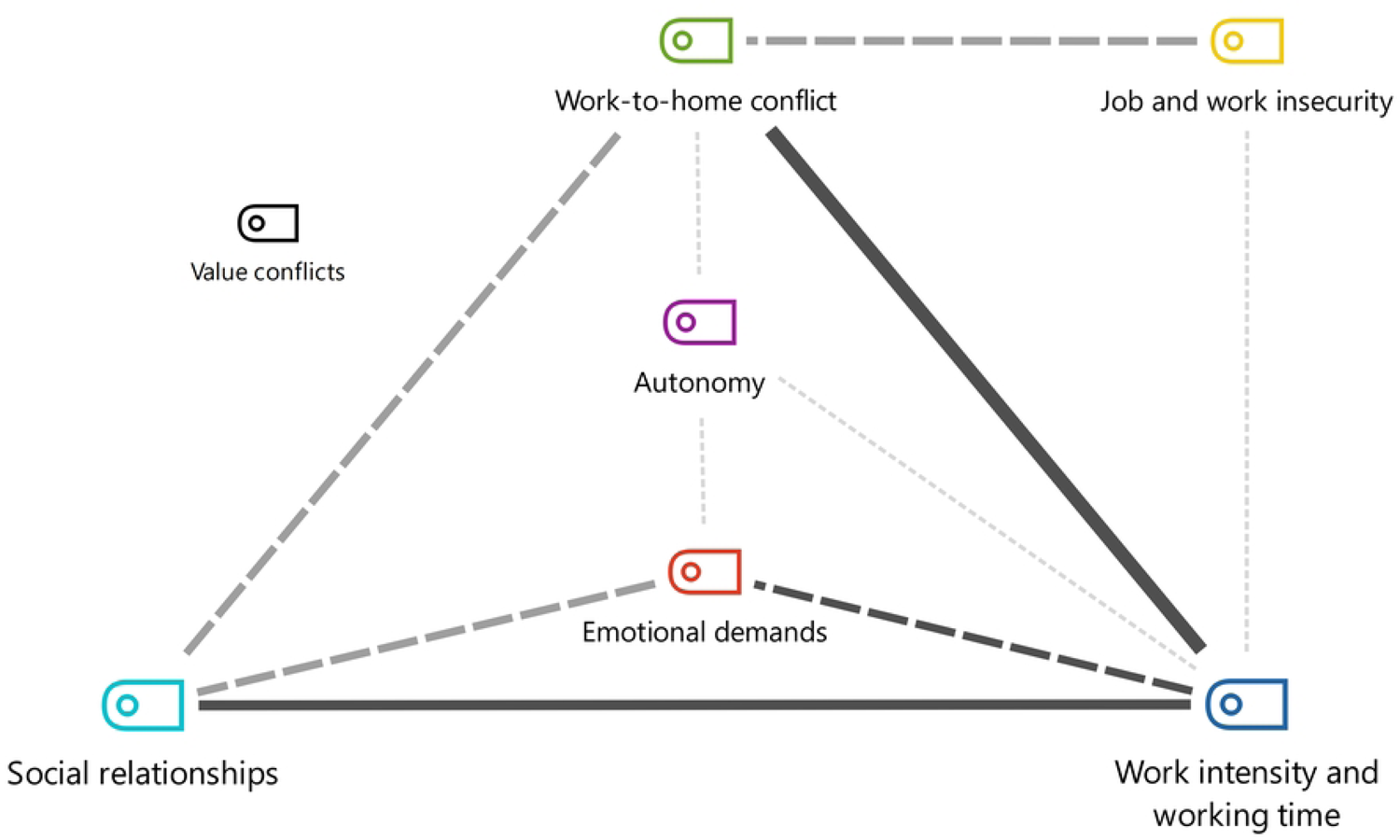
Countries of publication of articles included systematic review. Source: produced by authors.

Of the total articles selected, 22 were on part-time teleworking, 8 of which also addressed full-time teleworking, and 11 articles focused solely on full-time teleworking. A synthesis of all full articles was performed, providing positive and negative findings on interactions between work conditions and environment, organizational conditions, social conditions, job content and functions, efforts, besides individual and family characteristics of teleworkers. Following reading and analyses of the articles, these were classified into 7 categories (Table 3).

**Table 3.** Categories of psychosocial risk factors of work and number of articles

| Category | Number of articles |
| --- | --- |
| Work intensity | 14 |
| Emotional demands | 14 |
| Autonomy | 3 |
| Social relationships at work | 13 |
| Conflicts of values | 0 |
| Job insecurity | 3 |
| Home/work interface | 16 |

Source: produced by authors.

The analysis found that 18 articles addressed more than one category, with 8 addressing 2 categories, 8 investigating 3 categories and 2 involving 4 categories. A total of 15 articles explored one category only. However, none of the articles studied the 5^th^ category of “ conflict of values”.

The results from the MMAT are depicted in Fig 3. The qualitative synthesis of the articles reviewed, along with the main results for each publication including positive and negative aspects of teleworking, is presented in Table 4.

**Fig 3.**
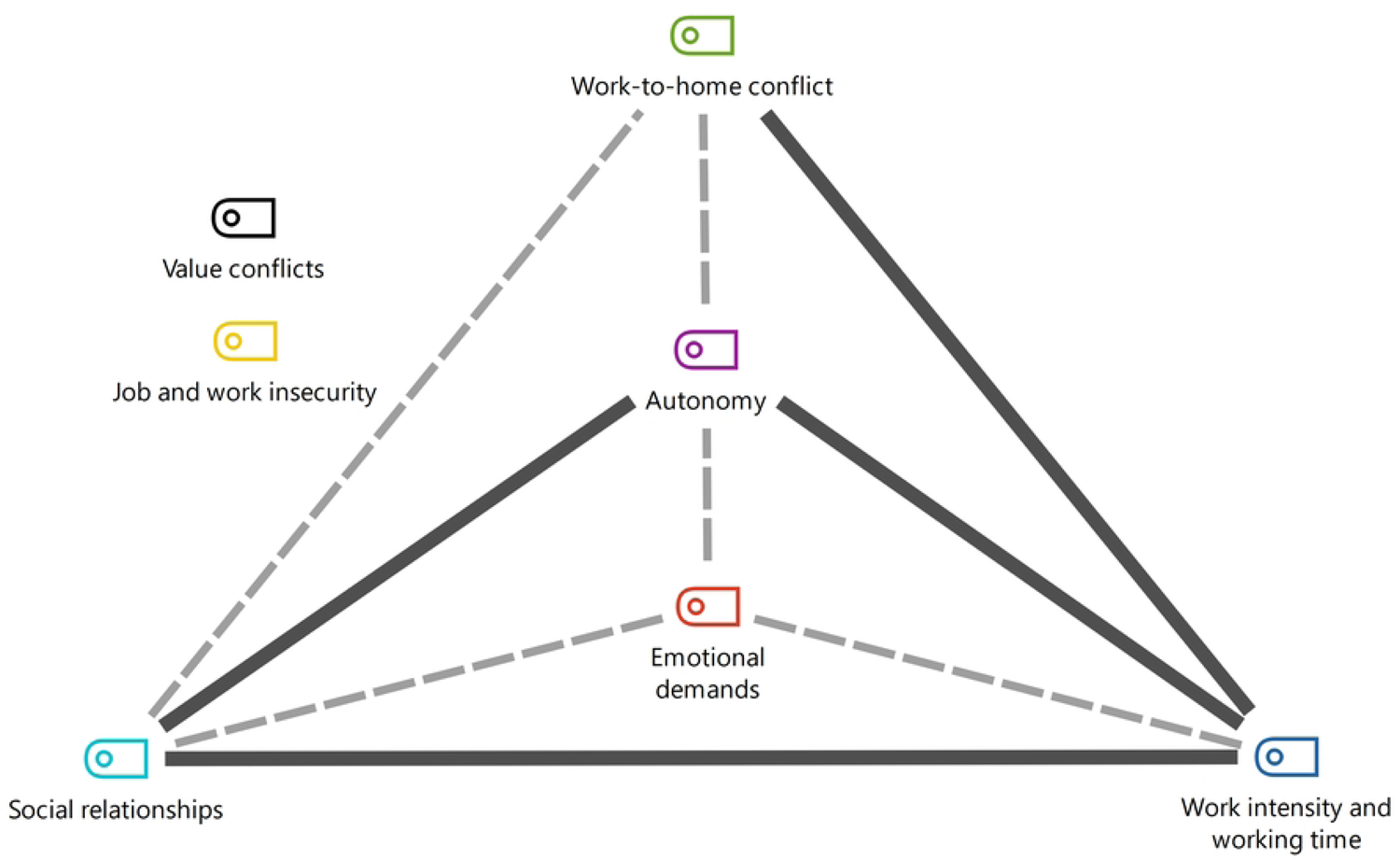
Results from Mixed Methods Appraisal Tool (MMAT). Source: produced by authors.

**Table 4.** Qualitative summary of articles reviewed.

| Author | Sample (demographics) | Results (factors of work investigated refer to interactions between environment and working conditions, organizational conditions, social conditions, functions and content of work, efforts, individual and family characteristics of teleworkers) |
| --- | --- | --- |
| Afonso, Fonseca and Teodoro, 2021 [36] | Portugal, alumni of Portuguese AESE Business (n=173)<br>**Full-time:** COVID-19 pandemic<br>**Data collection:** 2020 6-22 Aug (prior to full pandemic/face-to-face) | Quantitative – Descriptive cross-sectional<br>**Findings:**<br>Positive: Better sleep quality was associated with longer sleep duration and better job satisfaction.<br>Negative: High prevalence of poor sleep quality (74.1%) and general depression/anxiety (29.4%).<br>Male sex was negatively associated with perceived productivity. |
| Anderson, Kaplan and Vega, 2015 [37] | US, variety of government agency workers (n=102).<br>**Part-time & Full-time:** Mean 2.88 days per week<br>**Data collection:** 2014 | Quantitative<br>**Findings:**<br>Positive: Employees experienced more job-related positive affective well-being and less job-related negative affective well-being on days when they were teleworking compared to days they were working in the office. Teleworking can reduce level of emotions such as stress and anxiety and can increase emotions such as happiness and joy.<br>Negative: The affective consequences of telework seem to vary dramatically as a function of individual differences. |
| Barros and da Silva, 2010 [38] | Brazil, workers of the company Shell (n=15).<br>**Full-time:** None of the respondents chose to work home-office voluntarily.<br>**Data collection:** 2007 | Qualitative<br>**Findings:**<br>Positive: flexible working times; personal and family dimension, greater autonomy.<br>Negative: difficulty establishing limits for time dedicated to work. The company raised level of pressure to perform. Some pressures often appeared to be self-imposed. Difficulties regarding communication and interaction and lower visibility; difficulty for supervisors to monitor performance and establish a relationship of trust. Lack of synchronism; difficulties sharing information. Costs for career. Lack of boundaries between work and home life. Greater need to reconcile among women. |
| Bathini and Kandathil, 2019 [39] | India, 17 different companies (IT) (n=61).<br>**Part-time & Full-time:** 27 rarely/occasionally. 30 regularly.<br>**Data collection:** data not provided | Abductive<br>**Findings:**<br>Positive: reduces commute time<br>Negative: intensified work at home in exchange of telework. Portraying telework as an employee benefit or privilege helps management to orchestrate negotiation in their favor. |
| Belle, Burley and Long, 2015 [40] | US, workers from different sectors and organization types (n=10).<br>**Part-time & Full-time:** 4 telework/ 3-4 days per week and 6 full-time.<br>**Data collection:** 2013 | Phenomenological<br>**Findings:**<br>Positive: Expressions of organizational belonging included experiences that reflected self and other awareness, personal and professional fulfilment, support from others and participation.<br>Negative: Not belonging was apparent where there was a lack of credibility, conflict, a loss of stability and exclusion from ownership. |
| Bentley *et al.*, 2016 [41] | New Zealand, workers distributed across 28 organizations (n=804)<br>**Part-time:** 509 teleworkers/ 1-7 hours per week (low intensity) and 295 teleworkers/ ≥8 hours (hybrid).<br>**Data collection:** data not provided | Quantitative<br>**Findings:**<br>Positive: Organizational social support and teleworker support were associated with increased job satisfaction and reduced psychological strain. Organizational social support was regarded as having greater influence on satisfaction among low-intensity teleworkers.<br>Negative: Insufficient provision of organizational social support reduces job satisfaction and increases psychological strain due to the social isolation resulting from teleworking. |
| Biron and van Veldhoven, 2016 [42] | Netherlands, different occupations.<br>**Part-time:** 3 home days and 3 office days (n=77).<br>**Data collection:** data not provided | Multilevel analysis of mixed methods<br>**Findings:**<br>Positive: For workers with high work demands, ability to concentrate was higher and need for recovery lower, on home days than on office days.<br>Negative: on home days, generally high level of worktime control amplified the association between job demands and need for recovery—whereas this association was reversed when worktime control was generally moderate. |
| Cernas-Ortiz, Wai-Kwan, Cernas-Ortiz and Wai-Kwan, 2021 [43] | Mexico: Different occupations (n=214).<br>**Full-time:** Full-time during pandemic - face-to-face for 56% of sample pre-pandemic – 1<sup>st</sup> experience in teleworking.<br>**Data collection:** 2020 | Correlational. Non-experimental and cross-sectional.<br>**Findings:**<br>Positive: relationship between social connectedness outside of work and job satisfaction was positive and mediated by positive affective well-being, but not by reduction in negative affective well-being.<br>Negative: teleworking is not for everyone; it can cause social isolation and low job satisfaction. |
| Choi (2020) [44] | US, 143 subagencies of 15 Departments of executive power (n=428).<br>**Part-time (1-3 days per month) & non-telework/**<br>**Data collection:** 2011-2013 | Longitudinal analysis<br>**Findings:**<br>Positive: agencies that were more supportive and those with more teleworkers reported less voluntary turnover (0.02% in following year).<br>Negative: Organizational characteristics including average pay and length of service, and the proportions of different occupational categories, full-time employment, and women also had significant effects on voluntary turnover of employees |
| Darouei and Pluut, 2021 [45] | Netherlands, different occupations (such as legal sector, academia, IT) (n=34).<br>**Part-time** 2.7 days on average<br>**Data collection:** no data – performed prior to pandemic | Intraindividual model with daily surveys<br>**Findings:**<br>Positive: less time pressure, lower levels of work-family conflict on day. Family-work conflicts predicted individuals' next morning engagement and exhaustion levels and affective states towards the organization they worked for.<br>Negative: spiral of losses when experiencing work-family conflict. Negative effect on organization they worked for when work interfered with family life the previous day. |
| De Vries, Tummers and Bekkers, 2019 [46] | Netherlands, public servants (n=61).<br>**Part-time & Full-time**<br>**Data collection:** 2016 | Cross-sectional<br>**Findings:**<br>Positive: working from home did not affect work engagement. Leader-member exchange reduced impact of teleworking on professional isolation.<br>Negative: public servants experienced quite negative effects of teleworking, including greater professional isolation and lower organizational commitment on the days they worked entirely from home. |
| Duxbury & Halinski, 2014 [47] | Canada, 87 employees of medium-to-large public, private and NGO sectors (n=1806)<br>**Part-time** 1h-60h per week (Mean result 11.09h SD 13.5).<br>**Data collection:** 2011 and 2012 | Quantitative – hierarchical multiple regression analysis<br>**Findings:**<br>Positive: the telework arrangement seemed to help employees with higher work demands cope with these demands (i.e. work more hours with no concomitant increase in role overload).<br>Negative: the control offered by telework is domain specific (helps employees meet demands at work but not at home). Teleworking is not such an advantage to employees with lower work demands (overload increases faster). |
| Fukushima et al., 2021 [48] | Japan, living in Tokyo Metropolitan Area (n=1239)<br>**Part-time & Full-time**<br>Tele groups 1%-25%, 26%-50%, 51%-75%, and 76%-100% and other group not engaged in home-working<br>**Data collection: July and August 2020.** | Quantitative; cross-sectional<br>**Findings:**<br>Negative: Workers who teleworked were less physically active and had longer sedentary time during work time than those who worked at workplaces. Environmental factors of homeworking differed, e.g. size and layout of work space. |
| García-Salirrosas and Sánchez-Poma, 2020 [49] | Peru, university teachers (n=110)<br>**Full-time**<br>**Data collection: 2020** | Quantitative, cross-sectional study.<br>**Findings:**<br>Negative: A high prevalence of musculoskeletal disorders was found in the university teachers studied, particularly in the lumbar-dorsal spine and neck, and these disorders were associated with ergonomic risk factors such as poor posture and long working hours. |
| Hadi, Bakker and Häusser, 2021 [50] | Germany, workers from different professions (n= 178)<br>**Full-time:** mean 35.81h per week<br>**Data collection: April 2020** | Quantitative – multilevel analysis.<br>**Findings:**<br>Positive: Leisure can serve as an effective strategy to counterbalance emotional exhaustion.<br>Negative: Job and home demands (and COVID-19-related rumination) were associated with emotional exhaustion during teleworking in the COVID-19 pandemic. Daily home demands were the strongest predictors of emotional exhaustion. Emotional exhaustion caused by high demands impaired both individual well-being and also economic results (job performance). |
| Henke et al., 2016 [51] | US, active workers (n=3703).<br>**Part-time & non-teleworking/**<br>non-telework; telework outside working hours (50% or less of working time) and telework prime times (working hours 6-18h - 51% or more). low intensity (8 hours), medium intensity (9-32 hours), high intensity (33-72 hours), and very high intensity (73 hours).<br>**Data collection: 2010 and 2011** | Longitudinal analysis.<br>**Findings:**<br>Positive: Employees who telecommuted ≤8 hours per month were significantly less likely than nontelecommuters to experience depression. There was no association between telecommuting and stress or nutrition.<br>Negative: Telecommuting health risks varied by telecommuting intensity. The more teleworking, the higher the risk for stress. Nontelecommuters were at greater risk for obesity, alcohol abuse, physical inactivity, and tobacco use, and were at greater overall risk than at least one of the telecommuting groups. |
| Hoornweg, Peters and Van Der Heijden, 2016 [22] | Netherlands, bank organization (n=111).<br>**Part-time** ≤8h versus ≥8h per week - Mean 5.23.<br>**Data collection:** data not provided | Quantitative<br>**Findings:**<br>Positive: low telework intensity had no effect on individual productivity. The higher an employee was intrinsically motivated, the greater their productivity. High telework intensity combined with larger number of office hours can positively influence intrinsic motivation of employees.<br>Negative: the higher (> 8 hours per week) the telework intensity, the lower the individual productivity. |
| Kaduk, Genadek, Kelly and Moen, 2019 [52] | US, IT workers from 207 teams (n=758).<br>**Part-time** 20% of weekly hours corresponding to 1 day per week at home<br>**Data collection:** 2019 | Quantitative<br>**Findings:**<br>Positive: voluntary remote work was protective and more common. Employees working at least 20% of their hours at home and reporting moderate or high choice over where they worked had lower stress and intentions to leave the firm.<br>Negative: Involuntary variable schedules were associated with greater work-to-family conflict, stress, burnout, turnover intentions and other factors. Those engaged in substantial involuntary remote working reported greater insecurity at work than others. Women reported greater work-family conflicts, stress and anguish, but did not differ from men in terms of satisfaction at work, intention to leave or burnout. |
| Kelliher and Anderson, 2010 [53] | UK, 3 organizations - 15 IT personnel, 9 from pharmacies and 13 from consulting firms (n=37).<br>Quanti 729 remote workers and 228 on reduced hours contracts (n=2066).<br>**Part-time Full-time** all worked from home (typically 1 day); 14 full-time remote workers.<br>**Data collection:** data not provided | Qualitative and quantitative<br>**Findings:**<br>Positive: Flexible workers record higher levels of job satisfaction and organizational commitment than their non-flexible counterparts.<br>Negative: work intensification is being experienced by both those who work reduced hours and those who work remotely. |
| Koh, Allen & Zafar, 2013 [54] | Singapore, Survey<br>Singapore: Public Service - employees (56.5% executives junior or graduates); 27% management support), 15.8% corporate support/clerical, 0.5% manual workers (n=15,910)<br>**Part-time** 3 or more days per week; 1-2 days a week; 1-2 days a month; infrequently; not teleworking.<br>**Data collection:** 2011 | Quantitative<br>**Findings:**<br>Positive: teleworking users and non-users by choice reported higher levels of work-life balance than non-voluntary nonusers (not approved).<br>Negative: non-users due to non-authorization (not approved) or whose jobs were not conducive to teleworking reported a lower level of work-life balance. |
| Kumar, Kumar, Aggarwal and Yeap, 2021 [55] | India, public and private organizations (n=433).<br>**Full-time**<br>**Data collection:** May 2020: | Quantitative<br>**Findings:**<br>Positive: Role overload and change in lifestyle choices did not significantly affect job performance.<br>Negative: Family distraction, occupational discomfort and distress were significant in impacting job performance, with distress proving the most significant. During the COVID-19 pandemic, life satisfaction reduced due to a significant increase in distress levels and lowered job performances. |
| Lee and Kim, 2018 [56] | US, cabinets and independent agencies (n=194,739).<br>**Part-time** 3 or more days a week (3.27%), 1 or 2 days a week (11.45%), 1 or 2x per month (6.13%), very infrequently (16.1%), the remainder did not telework for different reasons. Total participation in some form of telework 71,951 (36.95%).<br>**Data collection:** 2011 | Quasi-experimental<br>**Findings:**<br>Positive: employees eligible to telework reported higher levels of perceived fairness, job satisfaction, and intention to stay than non-eligible employees.<br>Negative: employees who do not telework due to lack of technical or managerial support report significantly lower levels of perceived job fairness, job satisfaction and intention to stay than employees who telework. |
| Merisalo, Makkonen and Inkinen, 2013 [57] | Finland, creative and knowledge workers (n=971).<br>**Part-time & non-teleworkers/** 33 (4%) home teleworkers, 180 mobile or part-time teleworkers and 637 non-teleworkers. Full-time (+3 days), part-time (telecommuting), non-teleworkers<br>**Data collection:** 2010 | Quantitative.<br>**Findings:**<br>Positive: no difference was found in terms of knowledge intensity, creativity and e-capital between “home-anchored” workers and mobile or part-time teleworkers.<br>Negative: the results revealed the complexity of telework in both a theoretical and empirical context. |
| Müller and Niessen, 2019 [58] | Germany, teleworkers from different sectors (IT, production and manufacture, health and financial services, and others). **Part-time** working at least 20h a week at home at least 2 days a month. (n=195). Part-time employees (18%) Full-time employees (82%). <u>Part-time telework:</u> 29.81% of workweek homeworking (SD = 18.70; range: 1–90%) and 64.6% stated having 1 fixed day per week.<br>**Data collection:** data not provided | Quantitative – multilevel analysis<br>**Findings:**<br>Positive: part-time teleworkers reported higher use of self-reward, self-goal setting, and visualization of successful performance on home days than on office days. There were no indirect effects of working location on ego depletion through self-leadership. On home days, part-time teleworkers were more satisfied with their job at the end of the workday through self-goal setting. |
| Nguyen, 2021 [59] | Vietnam, undergraduates of the University of Transport and Communications and different professions (recruit. Facebook) (n=355)<br>**Full-time**<br>**Data collection:** 2020 April | Quantitative<br>**Findings:**<br>Positive: perception of home-based teleworking was positive with regard to fear of COVID-19. The presence of more than one child positively affected the attitude toward establishing the hybrid work mechanism.<br>Negative: difficulties in focusing on work and accessing data were negative factors. The presence of more than one child negatively affected the perception of telework. |
| Rodríguez-Modroño and López-Igual, 2021 [60] | EU28 countries, Sixth European Working Conditions Survey (n=35765).<br>**Part-time:** a) regular home-based teleworker - several x per month; b) highly mobile teleworker-ICT several times a week in at least 2 different places other than employer site; c) occasional teleworker- less frequent working from home/or other sites. **Data collection:** 2015 | Quantitative<br>**Findings:**<br>Positive: Occasional teleworkers were the group with the best job quality. Home-based teleworkers, especially women, presented better results than highly mobile workers in terms of working time quality and intensity. Negative: Highly mobile teleworkers were those with the worst job quality and work-life balance. Home-based teleworkers, especially women, had lower skills and discretion, income, and career prospects compared to highly mobile workers. |
| Shockley, Clark, Dodd, and King, 2021 [61] | US. Couples with 2 young children (<6 years), full-time workers (>32h) engaged in remote work or otherwise – 1st phase (n=274) and 2<sup>nd</sup> sample (n=179). **Part-time & Full-time** remote full-time at home; not engaged in telework; part-time remote (stable rota schedule); alternating remote working with couple; occasional remote working. **Data collection:** 2020 March (beginning of pandemic) and May (reopening of schools and daycares) | Mixed -qualitative and quantitative<br>**Findings:**<br>Positive: performance was better whenever there was some form of support from husband than when remote wife did all. The Alternating Days egalitarian category emerged as the overall strategy that best preserved wives' and husbands' well-being while allowing both to maintain adequate job performance. Negative: There was a lower performance for the *Remote Mini-shifts* and *Remote Need-Based Alternating models*. women in the *Remote Wife Does It All* class had the lowest well-being and performance. |
| Suh and Lee, 2017 [62] | South Korea. Teleworkers from 2 global IT companies (n=258)<br>**Part-time & Full-time** 104 high-intensity and 154 low intensity with > 2.5 days a week classified as high intensity and ≤ 2.5 days as low intensity telework. **Data collection:** 2015 | Quantitative<br>**Findings:**<br>Positive: Invasion of privacy and task interdependence did not have a significant effect in the high-intensity telework group. Negative: technostress reduced job satisfaction. Low-intensity teleworkers were more vulnerable to technologies than high-intensity teleworkers. The presence of information technology increased invasion of privacy in the low-intensity group. Task interdependence increased overload in the low-intensity group. The influence of job overload on strain was significantly greater in the low-intensity group. The influence of role ambiguity on strain was significantly greater in the high-intensity group. |
| Villavicencio-Ayub et al., 2021 [63] | Mexico. Different workers (n=724)<br>**Full-time**<br>**Data collection:** 2020 | Quantitative<br>**Findings:**<br>Positive: leadership, organizational support and education were fundamental aspects for satisfactory performance of the job. Negative: fear of infection, family conflicts and failure to follow usual patterns of life were associated with fear. Worry and economic problems emerged as factors that can negatively influence productivity. |
| Virick, DaSilva and Arrington, 2010 [64] | US, global telecommunications company (n=575)<br>**Part-time & Full-time** 85 teleworkers<br>**Data collection:** 2000 | Quantitative<br>**Findings:**<br>Positive: those with high drive and low enjoyment were more satisfied with high or low levels of teleworking, while all others preferred a moderate level of teleworking.<br>Negative: there may be a threshold in the number of days per week an individual can telecommute, beyond which the benefits to job satisfaction cease to continue to accrue. |
| Windeler, Chudoba and Sundrup, 2017 [65] | US. Study 1 – IT financial services (n=51). Study 2 - variety of industries (n=258).<br>**Part-time** Study 1) 1-2 x a week for 4 months Study 2) 98 regular practice and 160 1-2 x per month or less.<br>**Data collection:** 2016 | Quantitative.<br>**Findings:**<br>Positive: Practice of part-time telework can attenuate the relationship between interpersonal interaction and work exhaustion.<br>Negative: work exhaustion increased as interpersonal interaction increased. Interdependence increased work exhaustion for teleworkers and non-teleworkers. |
| Yoshimoto et al., 2021 [66] | Japan, variety of areas (n=1941)<br>**Part-time & Full-time**<br>**Data collection:** During second wave in Japan. Jul/Aug 2020 | Cross-sectional<br>**Findings:**<br>Negative: workers reported that their pain worsened during the COVID-19 pandemic. Telework, physical activity and psychological stress were significantly associated with pain augmentation. |
| Zhang, Moeckel, Moreno, Shuai and Gao, 2020 [67] | Germany (n=188,081)<br>**Part-time & Full-time** 12.85% infrequent/frequent, sometimes 74.15% (< 50%) and frequent (50-100%).<br>**Data collection:** 2010 | Quantitative<br>**Findings:**<br>Negative: the presence of children increased work-to-family conflict, family-to-work conflict and also triggered re-arrangement of housework within couples and aggravated gender differences. |

Source: produced by the authors.

## 4. DISCUSSION

### 4.1 Dimensions of psychosocial risk factors

The evidence on psychosocial risk factors at work is heterogeneous, given these are related to work organization and different variables. Therefore, the present study sought to shed light on the issue by identifying the amplified (most found) and reduced (least found) dimensions comparatively in the published articles, for part-time and full-time home-based teleworking (Table 5).

**Table 5.**
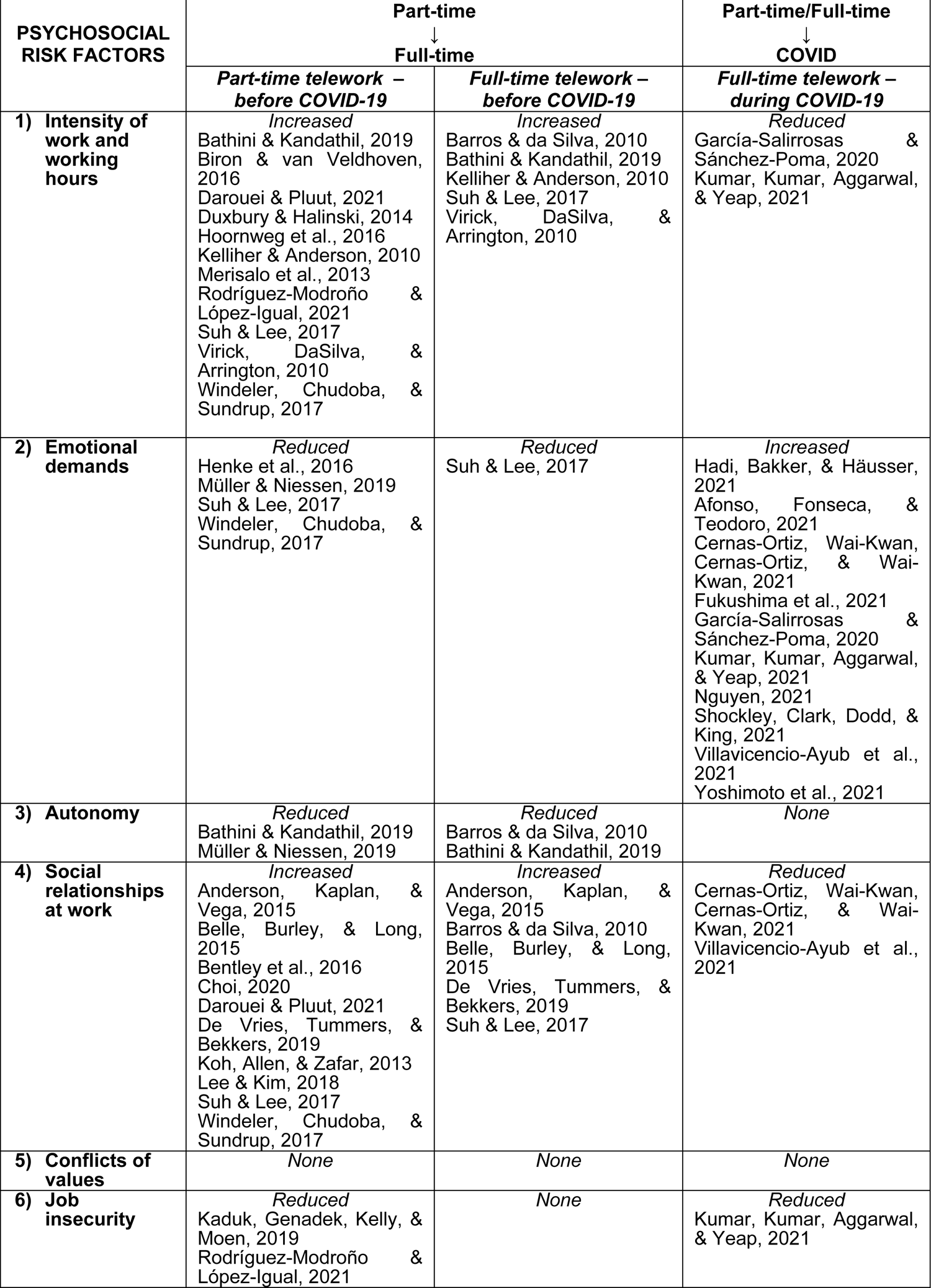
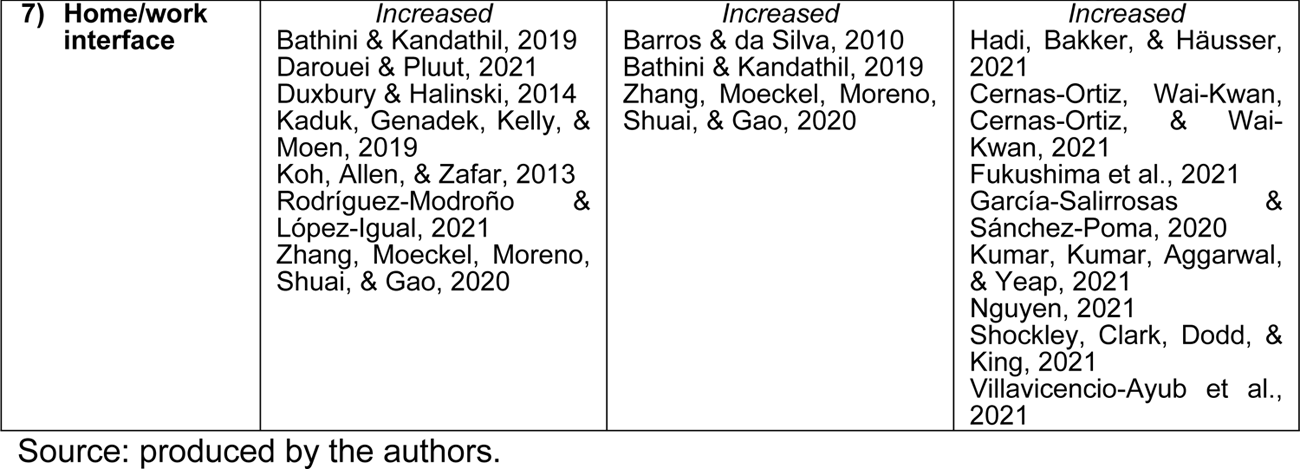
Analysis of results by authors related to the dimensions of the psychosocial risk factors found.

#### 4.1.1 Work intensity and working hours

Telework is generally associated with intensification of work [49,68,69,70]. Bathini and Kandathil [39] found that, in certain settings, managers use telework as a strategy to reduce reluctance of workers to undertake an intensified work schedule. Intensification of work is not a reciprocal exchange between employee and employer. Telework can also be a facilitator of longer working days, making it easier for the employer to demand work hours that exceed limits stipulated by law [39]. In exchange for greater autonomy, companies also raise the level of pressure: somebody from the company sends an email and expects a rapid response, obliging the individual to reorganize their priorities, exceeding working times and expecting more from the employee [38].

The phenomenon of “unimposed” intensification is highlighted by Kelliher and Anderson [53], in which may remote workers showed that when home-working, the working day tended to be longer, with more intense effort made. The only study conducted prior to the pandemic involving solely full-time teleworkers outlined the difficulties establishing limits for time dedicated to work [38].

On the other hand, authors showed that home-based teleworking can be associated with more positive emotional results [42,45,71]. Flexible hours are also an aspect perceived as positive by individuals [38]. However, when the working hours are controlled by the company, these positive results may not persist [42].

Intensification of work can be associated with autonomy. According to Duxbury and Halinski [47], the control over work demands afforded by telework becomes more important as working hours per week rise.

Studies carried out during the pandemic confirm intensification of home-based full-time teleworking, both through longer work hours in front of the computer during the day, and working weekends, sometimes under pressure from superiors [49, 55].

On the other hand, the role of leaders can favor the productivity attained at home, which can be both higher and more enjoyable [63]. However, the same authors showed that the lack of regulations governing home-working precludes the establishment of a work-life balance, clashing with productivity and working hours [63].

Productivity can be viewed as a paradox in that its increase can be related to the higher number of hours worked beyond the normal working week owing to the lack of regulation or control of times worked. Hoornweg, Peters and Van Der Heijden [22] examined the productivity-teleworking relationship, concluding that the higher (> 8 hours per week) the telework intensity, the lower the individual productivity.

Work intensity and working hours are associated with job complexity in its quantitative and qualitative dimensions [28]. This has drawn the attention of international organs with respect to health and safety at work, working hours, life-work balance, occupational health and safety risk prevention in teleworking [69]. Part of this advance in the discussions, regulations and implementation includes the “right to disconnect” [69].

#### 4.1.2 Emotional demands

This dimension was not prominent prior to the COVID-19 pandemic. Barros and da Silva [38] found that pressure to deliver was often self-imposed, driven by the feeling that responsibility for results rests with the individual. These findings are congruent with the study by Müller and Niessen [58], in which employees reported higher use of self-reward, self-goal setting, and visualization of successful performance on home days. These results may be explained by the social and professional isolation promoted by teleworking, the worker centrality, individual performance, and loss of group working/learning.

Suh and Lee [62] reported feelings of invasion of privacy among teleworkers due to the presence of technology. Windeler, Chudoba and Sundrup [65] found that external interaction (interpersonal) increased work exhaustion for part-time teleworkers.

The intensification of emotional demands during the COVID-19 period impacted job performance [50,55,63]. Effects such as social isolation, high exposure to electronics use and fear can form a backdrop for risk factors and impact on the mental and physical health of workers, such as emotional exhaustion, symptoms of anxiety and depression and augmented pain [36,48–50,66]. Fear of COVID-19 was associated with those who most liked to adhere to home-based teleworking [59].

Gender issues were hitherto little explored in studies on teleworking. Some findings are conflicting, such as those of Afonso, Fonseca and Teodoro [36] showing that men reported lower rates of productivity than women in teleworking versus Shockley et al. [61] who found that productivity was affected in both men and women with children during the pandemic.

Married professionals had higher work load due to COVID-19 and high risk of distress [55]. The relationship of family distractions, lifestyle choice, role overload and operational discomfort with life satisfaction, respectively, was sequential and positively mediated by distress and job performance [55].

#### 4.1.3 Autonomy

This is defined as the possibility of the worker being active as opposed to passive with regard to their job, participation in production and leading their professional life [28].

This aspect was addressed in two pre-COVID studies, where Bathini & Kandathil [68] noted that control of time autonomy was a dominant paradox, in as far as workers who enjoyed job autonomy through use of flexible work arrangements ultimately worked more and for longer.

Müller and Niessen [58] examined the mediating role of autonomy in part-time teleworkers. Autonomy plays a mediating role in the association between working location and self-reward, self-goal setting, visualization of successful performance and evaluation of beliefs and assumptions [58].

A group of researchers that analyzed psychosocial risks of work recommended that further studies (special review of literature review, specific epidemiological studies, qualitative studies) be conducted to examine the effects of worker autonomy when the intensity of other psychosocial risk factors are very high [28]. It was clear that, when telework was mostly voluntary during the pre-COVID era, autonomy was a pre-requisite. However, given the compulsory nature of telework during the pandemic, further investigations into this dimension were called for.

#### 4.1.4 Social relationships at work

Unlike emotional demands, the dimension of social relationships at work was more explored in pre-pandemic studies. Social and professional isolation is one of the most reported disadvantages in teleworking - a model that individualizes life at work. Particularly, for mental health, and collective action to confront the guarantee of rights, this is a key critical point.

This aspect is associated with difficulties involving communication (synchronism), integration and lower visibility [38, 46]. However, this problem increases in employees engaged in higher intensity teleworking who work remotely for long periods, reducing job satisfaction and increasing psychological strain [41].

Working entirely from home led to the public servants studied by De Vries, Tummers and Bekkers [46] experiencing lower organizational commitment, an effect not seen on days worked partially from home. According to Belle, Burley and Long [40], not belonging among high-intensity teleworkers might be associated with a lack of credibility, conflict, a loss of stability and feeling of invisibility.

However, it is also important to observe the relationship between teleworkers and non-teleworkers. The findings of Suh and Lee [62] showed that task interdependence increased overload in the low-intensity teleworking group, but did not have a significant effect in the high-intensity telework group. In the study by Windeler et al. [65], interdependence increased work exhaustion for both teleworkers and non-teleworkers.

The findings of studies published during the pandemic echoed those of previous studies in which social support mechanisms, in and outside the workplace, are relevant for teleworker satisfaction [43]. Leadership, organizational support and education are also fundamental aspects for satisfactory job performance [63]. The fact workers of the companies were in the same situation may have promoted greater affect via social support during remote working between workers and supervisors.

#### 4.1.5 Conflicts of values

No evidence characterizing psychosocial risk factors related to conflict of values was found in this systematic review of the literature. Gollac & Bodier [28] showed that conflict of values rarely features in surveys on working conditions and on the nature of psychosocial risk factors. Conflicts of values can take on the form of an ethical conflict, when the worker is asked to act in breach of their professional, social or personal values, and also in other ways, such as being prevented from carrying out work of quality and the feeling of doing useless work through the tradeoff between quality and quantity [28].

The lack of relevant studies on this dimension in this systematic review does not imply the factor is non-existent. A recent study published in 2022 offered clues in an excerpt from a workeŕs narrative that pointed to the need for studies that address this category:

> Now, there is a feeling and concern on the surface that the field cannot be done very well and continuous development exhausts work communities. **This brings me as a developer an ethical conflict** and in a way eats away the motivation to do development work. I wonder how the development of work in everyday life a more natural part of the whole structure could be, neither detached, nor temporary, **nor a necessary evil imposed by the employer that tears at many different goals and objectives**. [72] (p.9, bolding by authors of present review).

#### 4.1.6 Job insecurity

In the United States of America (USA), full-time employees had a high level of commitment [44]. However, also in the USA, workers engaged in substantial involuntary remote working reported greater job insecurity than others [52].

Moreover, telework can reinforce traditional gender roles [60]. An analysis of the 28 countries of the European Union showed that women teleworking from home, besides receiving a 31% lower salary than men, also had greater perceived insecurity at work than men [60].

Amid the different governmental measures for tackling the pandemic, aspects of job insecurity were heightened. This manifested in the form of intensification of work to avoid job loss and accepting lower salaries [55, 63].

#### 4.1.7 Home/work interface

The family appears to act both as a source of distraction and guide for priorities [38]. The family does not always recognize that boundaries must be drawn separating work and home life, given it is easily presumed the individual is available at any time, in addition to issues of sharing spaces [38, 39]. Work-to family conflict can affect levels of commitment and affective states such as emotional exhaustion, job engagement, and negative affects toward the organization [45].

Zhang, Moeckel, Moreno, Shuai e Gao [67] suggested that, as one of the most important characteristics in the family sphere, children play a key role in telework. The presence of children not only increased work-to-family conflict but also triggered redivision of housework within couples and aggravated gender differences [47, 67].

For women, the need to reconcile is greater, because involvement in child care tends to be higher [38]. They also reported more work-to-family conflict, stress and anguish, but did not differ from men in terms of job satisfaction work, intention to leave or burnout [52]. According to Rodríguez-Modroño and López-Igual [60], full-time home-based teleworkers, both men and women, had better conciliation between work and family and social commitments, whereas occasional teleworkers had worse professional balance.

During the pandemic, it was found that interpersonal relationships within the home were disrupted by confinement, leading to conflicts in personal and family life [63]. From a personal perspective, positive relationships were found between daily job and home demands, but unfavorable relationships with emotional exhaustion developed during teleworking [50]. Greater family distractions and role overload increased distress and negatively impacted job performance [55].

Corroborating pre-pandemic studies, child care fell mostly to wives. Husbands helped occasionally, for example, one day a week or during a wifés important meeting [61].

### 4.2 Inter-relationship between dimensions of psychosocial risk factors

Psychosocial risk factors at work stem from interaction of a social situation (organization, status, economic conditions, work conditions) with mental functioning [28].

Co-occurrence analysis, i.e. frequency with which dimensions appear together in the same study, resulted in the different intersections depicted in Figs 4, 5, and 6. The dimensions exhibited strong, medium or low connection with each other, according to the frequency of telework practiced.

**Fig 4.**
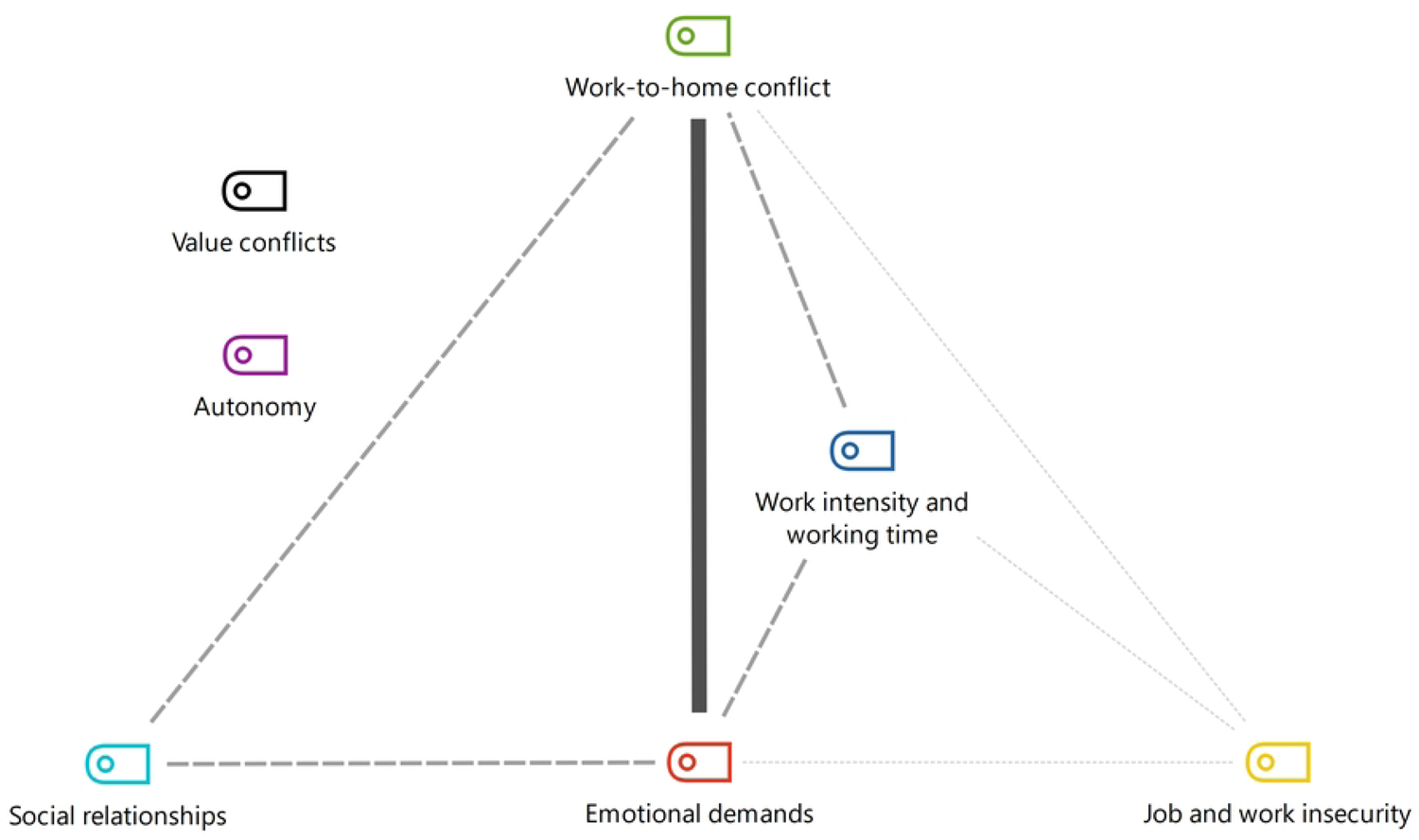
Co-occurrence of dimensions of part-time teleworking before pandemic. Source: produced by authors.

**Fig 5.**
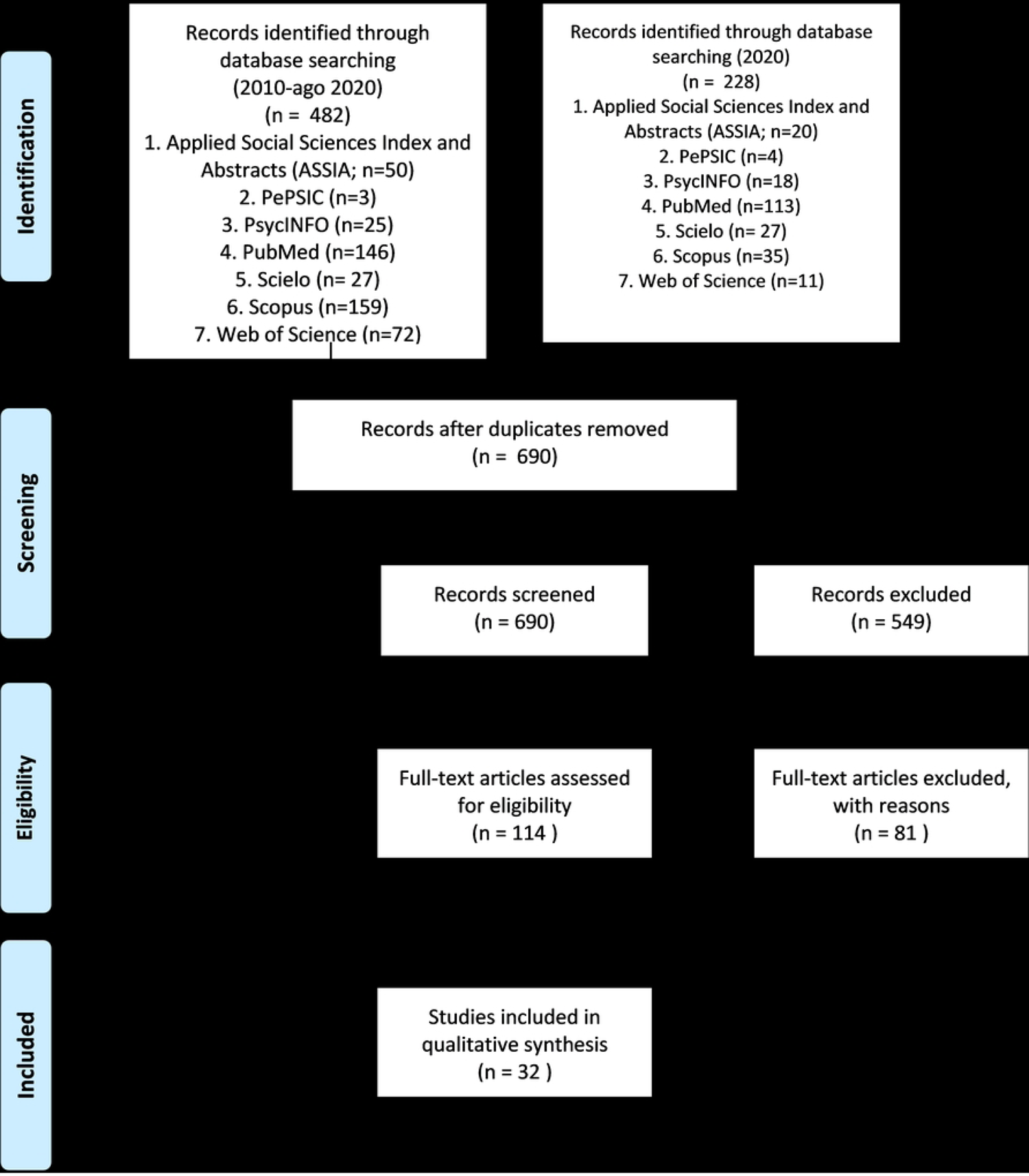
Co-occurrence of dimensions of full-time teleworking before pandemic. Source: produced by authors.

With regard to co-occurrence of the dimensions of part-time teleworking before the pandemic, a strong connection was identified for work intensity and working time with social relationships and work-to-home conflict (Fig 4) [39,45,47,54,60,62,65]. The results cited corroborate the fact that the dimensions studied do not manifest individually.

Results also revealed that the strong relationships of part-time teleworking persisted for full-time teleworking before the pandemic, on top of the strong association of the autonomy dimension (Fig 5) [38,39,62]. These results suggest that, before the pandemic, autonomy was considered a pre-requisite for practicing full-time teleworking.

For part/full-time teleworking during the pandemic, the previous inter-relationships became moderate or weak. However, the main co-occurrence took place between emotional domains and home-to-work conflict outlined earlier [43,48–50,55,59,61,63].

It should be noted that the emotional demands found are not related to clients and external relationships. In this case, they were associated with distress during the COVID-10 period during isolation, included by the authors in the classification of Gollac & Bodier [28]. Another change occurred in as far as autonomy was no longer highlighted teleworking, given that this became compulsory.

However, despite the potential risks, it should be noted that many variables are ambiguous, being positive or negative. These results are presented in the qualitative synthesis of the articles (Table 4) and in the Discussion section.

Telework presents positive elements highlighted by workers, such as the reduction in commuting time, greater freedom of work schedules, improved mealtimes. However, it is important to note that it can significantly deepen unequal relationships between workers and employers, in which what is lost is always much greater than what is gained.

On the other hand, the greater control of time by the worker may be illusory. Due to greater demands of work, there is no other way to achieve the goals and demands, unless extending his teleworking day. Before the pandemic, studies have already indicated that the experience of temporal and spatial reorganization of the work at home commonly lead to an intensification of work, since the command and control over time pass to the workers [73–75].

Therefore, flexible working hours and teleworking have become a synthesis of work intensification, which we can characterize as a contradiction to the search for work-life balance [18].

There is a share of conflicts that are experienced internally by workers - Whom to preferably attend to? Family? Work? The temporal references of time at work and time off work have dissolved significantly. These aspects are not mentioned in the literature discussed here.

The dimensions of psychosocial risk factors are not alone and may act as protective factors in seeking balance and prevention of risk. The models depicted in Figs 4, 5 and 6 contribute with points that can be prioritized in a system for assessment, monitoring and intervention in the psychosocial risks of teleworking.

**Fig 6.**
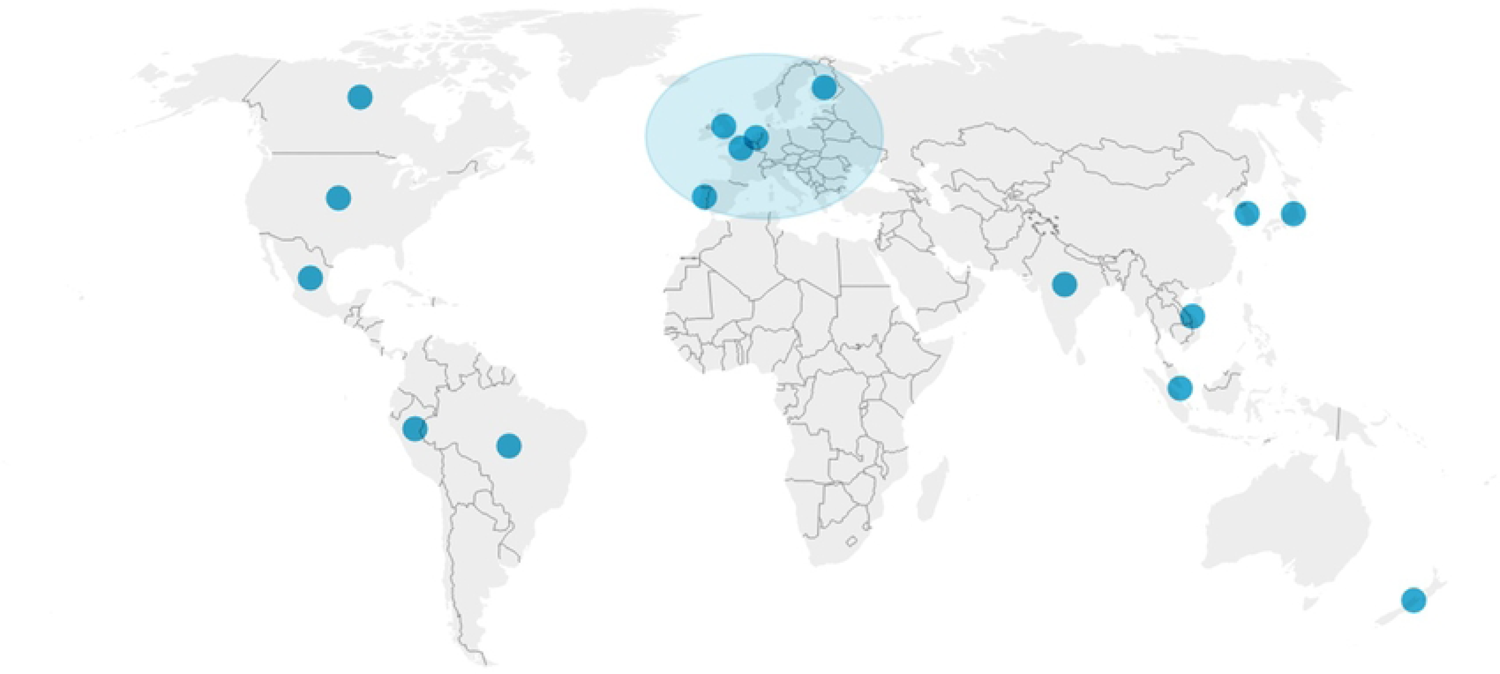
Co-occurrence of dimensions of full/part-time teleworking during pandemic. Source: produced by authors.

### FINAL CONSIDERATIONS

The systematizing of the psychosocial risk factors of part and full-time teleworking did not allow metrics to be determined but did provide pointers on their differences, where these were impacted by the current health crisis. This entails analyzing job complexity, different professions, working conditions, countries and legislation.

We suggest that the benefits reported by studies on teleworking be interpreted with caution. Teleworking is not devoid of risks and it is important to consider the specificity and complexity of each activity undertaken. However, the dearth of longitudinal studies precludes any meaningful predictions of their long-term impacts.

Of the 7 dimensions of psychosocial risk factors, more than one was identified in some of the studies reviewed. Before the pandemic, research attention was centered on dimensions involving work intensity and working times and social relationships at work, particularly for part-time teleworking. During the pandemic and full-time teleworking, emotional demands featured more prominently in studies. However, the home-work interface remained the central focus before and after the pandemic and during part and full-time teleworking.

The present review helps inform and foster debate on public policies addressing psychosocial risk factors. The study yields evidence on which to base the search for possible solutions. In addition to pooling qualitative, quantitative and mixed methods studies, the review highlights the significant increase in home-based teleworking and the psychosocial risk factors associates with this work modality.

### Limitations and recommendation for future studies

The goal of systematic reviews with qualitative evidence is not to assess the effects of the interventions. However, such mapping reviews can complement systematic reviews of efficacy studies, help explain and interpret results of syntheses of evidence on “what works”, as well as answer other questions, which can serve to inform policy and practice.

There is a pressing need to organize definitions and clarify concepts regarding teleworking. This can help determine the prevalence, frequency and intensity with which employees work from home or practice home-based teleworking, and also track changes in these parameters over time.

Future studies should focus on the data collection instruments employed and analyze categories not found (conflict of values) in the present systematic review. Lastly, longitudinal studies should be encouraged as valuable tools for analyzing public policies on teleworking.

## Additional information

This systematic review was part of a doctoral thesis on the implementation of telework in the Brazilian federal judicial sector. The study was conducted during a PhD program in Public Health under the research topic: Organization of production processes and workerś health, School of Public Health of the University of São Paulo, Brazil.

## Data Availability

The data will be kept in the public repository of theses of the University of São Paulo, Brazil because they are part of the thesis developed by the author Evelise Dias Antunes. https://www.teses.usp.br/ Also available in PROSPERO 2020 platform - International Prospective Register of Systematic Reviews (number CRD42020191455). https://www.crd.york.ac.uk/prospero/display_record.php?ID=CRD42020191455

## Acknowledgements

The authors would like to thank Dr Tânia Maria de Araújo and Dr. Arlete Ana Motter for their comments.

